# Prognostic factors for mortality, ICU, MIS-C and hospital admission due to SARS-CoV-2 in paediatric patients: A systematic review and meta-analysis

**DOI:** 10.1101/2024.02.23.23298451

**Authors:** Constantine I. Vardavas, Katerina Nikitara, Alexander G. Mathioudakis, Dimitris Delialis, Valia Marou, Nithya Ramesh, Kimon Stamatelopoulos, Georgios Georgiopoulos, Revati Phalkey, Jo Leonardi-Bee, Charlotte Deogan, Favelle Lamb, Aikaterini Mougkou, Anastasia Pharris, Jonathan E. Suk

## Abstract

**Background:** There is a paucity of data on the factors associated with severe COVID-19 disease, especially in children. This systematic review and meta-analysis aim to identify the risk factors for acute adverse outcomes of COVID-19 within paediatric populations, using the recruitment setting as a proxy of initial disease severity.

**Methods:** A systematic review and meta-analysis were performed representing published evidence from the start of the pandemic up to 14 February 2022. Our primary outcome was the identification of risk factors for adverse outcomes, stratified by recruitment setting (community, hospital). No geographical restrictions were imposed. The Grading of Recommendations Assessment, Development and Evaluation (GRADE) methodology was used to evaluate the certainty in the body of evidence for each meta-analysis. In anticipation of significant clinical and methodological heterogeneity in the meta-analyses, we fitted logistic regression models with random effects.

**Findings:** Our review identified 47 studies involving 94,210 paediatric cases of COVID-19. Infants up to 3 months were more likely to be hospitalised than older children. Gender and ethnicity were not associated with an increased likelihood of adverse outcomes among children within the community setting. Concerning comorbidities, having at least one pre-existing disease increased the odds of hospitalisation. Concerning BMI, underweight children and severely obese were noted to have an increased likelihood of hospital admission. The presence of metabolic disorders and children with underlying cardiovascular diseases, respiratory disorders, neuromuscular disorders and neurologic conditions were also more likely to be hospitalised. Concerning underlying comorbidities, paediatric hospitalised patients with congenital/genetic disease, those obese, with malignancy, cardiovascular diseases and respiratory disease were associated with higher odds of being admitted to ICU or ventilated.

**Interpretation:** Our findings suggest that age, male, gender, and paediatric comorbidities increased the likelihood of hospital and ICU admission. Obesity, malignancy, and respiratory and cardiovascular disorders were among the most important risk factors for hospital and ICU admission among children with COVID-19. The extent to which these factors were linked to actual severity or where the application of cautious preventive care is an area in which further research is needed.

## INTRODUCTION

Children had the lowest case notification rates in the early stages of the pandemic, though studies have since suggested this may have been related to increased frequencies of asymptomatic and mild illnesses in kids and lower testing rates rather than a decline in susceptibility (1). Children also seem to present, in principle, either no symptoms or have a milder clinical picture, leading to lower rates of hospitalisation or death compared to adults (3). Though the intensity or severity of the clinical manifestations of COVID-19 differs from those in adults in the short term, current data show that prolonged symptom duration also exists in children (4). However, in a small proportion of paediatric patients, severe complications could occur, as well as hospitalisations or even death (5).

Due to the novelty of SARS-CoV-2 and the rapid evolution of its variants, there is a paucity of data on the factors associated with severe COVID-19, especially in children, since many do not present with acute symptoms. This makes it imperative to identify the patient factors associated with severe COVID-19, which would indicate patient management pathways within the healthcare system and can be used to guide national decision-making bodies on COVID-19 mitigation strategies and may contribute to the evidence base for future respiratory pandemics. This systematic review and meta-analysis aims to identify the risk factors for adverse outcomes of COVID-19 within the paediatric population, using the recruitment setting (community vs. hospital) as a proxy of initial disease severity.

## METHODS

The systematic review adhered to PRISMA (Preferred Reporting Items for Systematic Reviews and Meta-Analysis) (6) and MOOSE (Meta-analyses Of Observational Studies in Epidemiology) guidelines (7). The protocol of this systematic review was pre-reviewed by the European Centre for Disease Prevention and Control (ECDC). The protocol was not pre-registered in any database for systematic reviews.

### Outcomes and Inclusion/exclusion criteria

Randomised controlled trials, non-randomised controlled trials, prospective and retrospective cohort studies, case-control studies, and analytical cross-sectional studies with no geographical limitation were considered eligible provided (i) they evaluated patients between the ages of 0-18 years old with clinically diagnosed or laboratory-confirmed COVID-19 in one of the following settings: community, hospital, intensive care unit (ICU); (ii) they assessed the associations between underlying conditions (as risk factors) and primary adverse outcomes of COVID-19. All underlying clinical conditions (risk factors) reported in the original studies were considered acceptable for data extraction.

### Screening and data extraction

Relevant peer-reviewed studies published in English were identified within Medline (OVID) and EMBASE (OVID) from the start of the pandemic until 14 February 2022. Subject headings relating to COVID-19 and epidemiological study design terms were used to develop a comprehensive search strategy presented in **Appendix 1**. Reference lists of all included studies and identified reviews were also screened to identify additional relevant studies. Systematic and non-systematic literature reviews were excluded, but their references were screened. Full texts of potentially eligible studies were evaluated independently by two reviewers. Disagreements or uncertainties in the screening stages were resolved through discussion and consensus.

An ad hoc designed structured form was used for the adjusted data from each eligible study, including details on the study design, baseline characteristics of the participants, risk factors and outcome data. For accuracy, two reviewers extracted each study’s data, and disagreements were discussed and resolved by a third reviewer. Adequate information was extracted to allow us to identify overlapping populations across studies; we prioritised including data from the study with the largest population and a more rigorously described methodology.

The methodological quality of each included study was evaluated independently by two reviewers using the Joanna Briggs Institute (JBI) standardised critical appraisal tool for the appropriate design (8). Disagreements were resolved with discussion and, when necessary, adjudication by a third reviewer. The results of the quality appraisal are presented in **Appendix 2**.

### Assessing the certainty of the evidence

The Grading of Recommendations Assessment, Development and Evaluation (GRADE) methodology was used for evaluating the certainty in the body of evidence for each meta-analysis (8). In line with GRADE recommendations for the assessment of the evidence of prognostic factors, we initially ascribed high certainty to all our findings. Studies were subsequently rated down according to several factors, including study limitations of the included studies, inconsistencies, indirectness or imprecision of the results, or evidence of publication bias. The level of certainty was rated up in cases of large observed effects.

### Statistical Analysis

In anticipation of significant clinical and methodological heterogeneity in the meta-analyses, we fitted logistic regression models with random effects. All variables were dichotomous and analysed as Odds Ratios (OR) with corresponding 95% confidence intervals (95% C.I.). Heterogeneity was quantified using the I2 statistics. We considered values above 75% to represent considerable heterogeneity. To facilitate the interpretability of the results and in line with recommendations by GRADE (14), we also present the absolute Risk Differences (RD) per 1,000 COVID-19 patients with corresponding 95% C.I. The median prevalence of the evaluated risk factors and the median incidence rate of each adverse outcome in each recruitment setting were used for calculating the absolute RDs.

## RESULTS

In the review, we included 47 studies involving 94,210 paediatric cases of COVID-19 (**PRISMA flowchart: Figure 1**). Recruitment dates ranged from 1 January 2020 to 24 September 2021 but were primarily based in 2020. There were 26 cohort studies (5, 9–33), 10 cross-sectional studies (34–43), seven chart reviews (44–50), three case-control studies (51–53) and one case series (54). Details on the characteristics of the included cohort studies are presented in **Table 1**.

**Figure 1.**
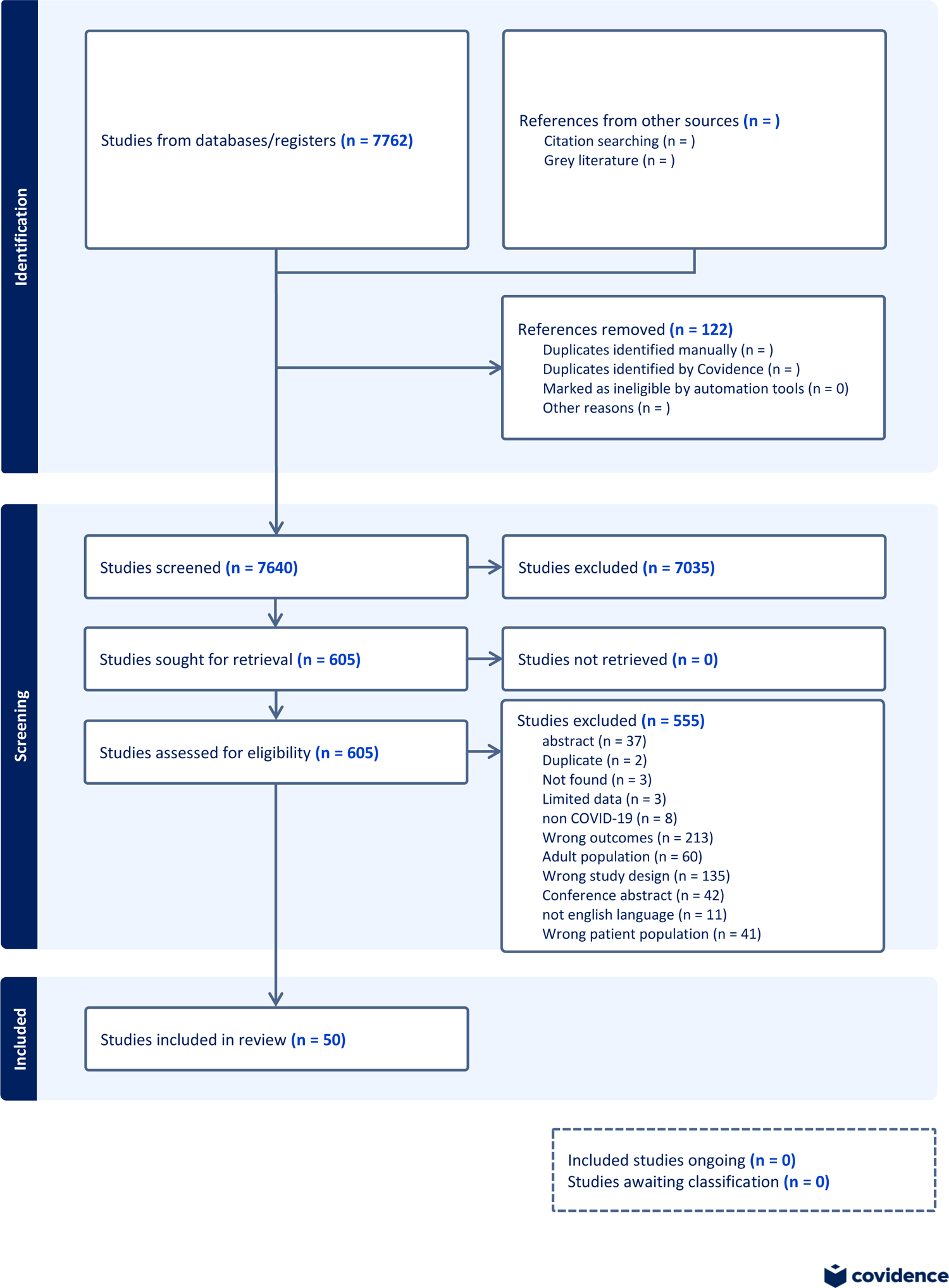
Flowchart1.

**Table 1.**
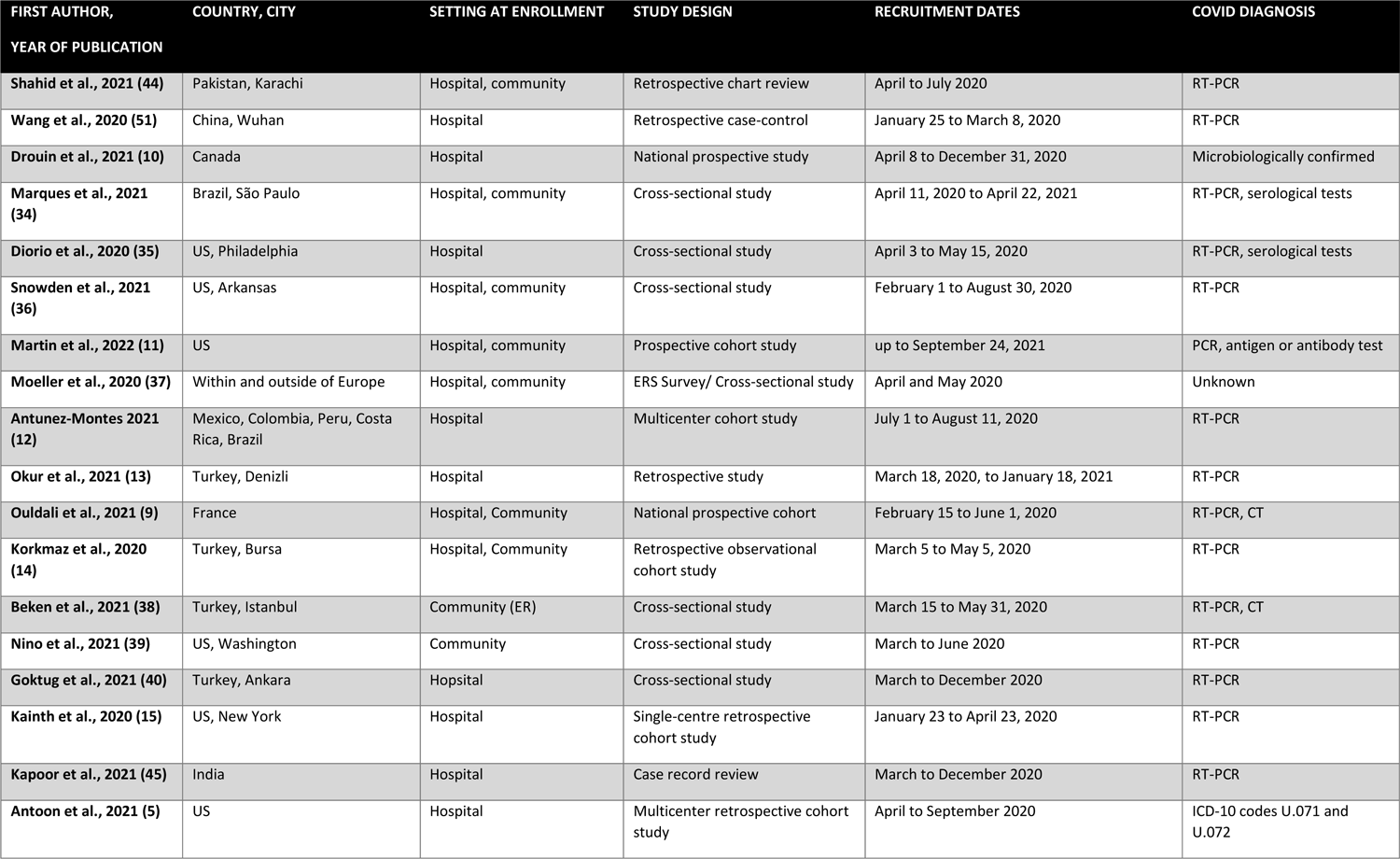
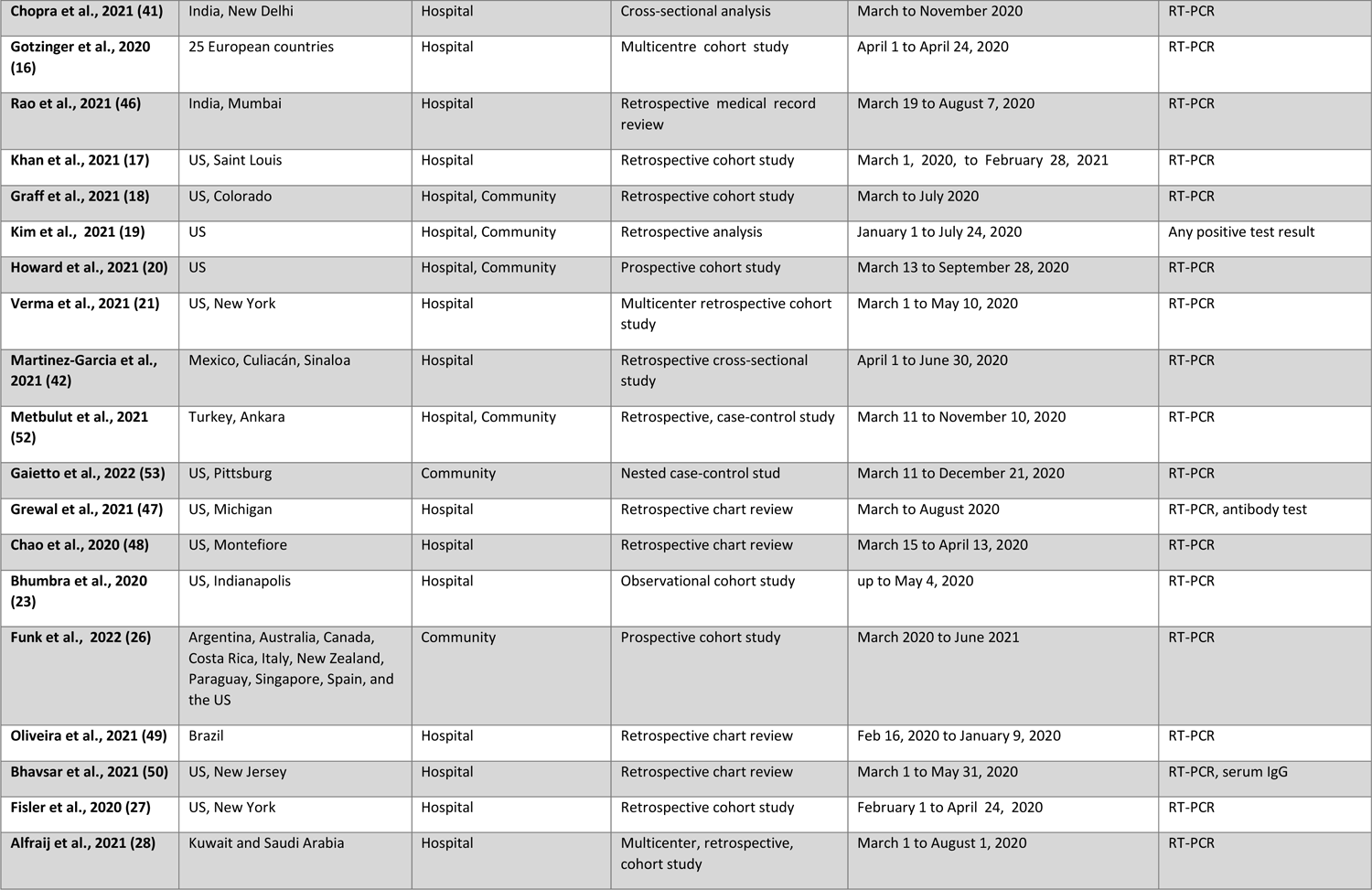
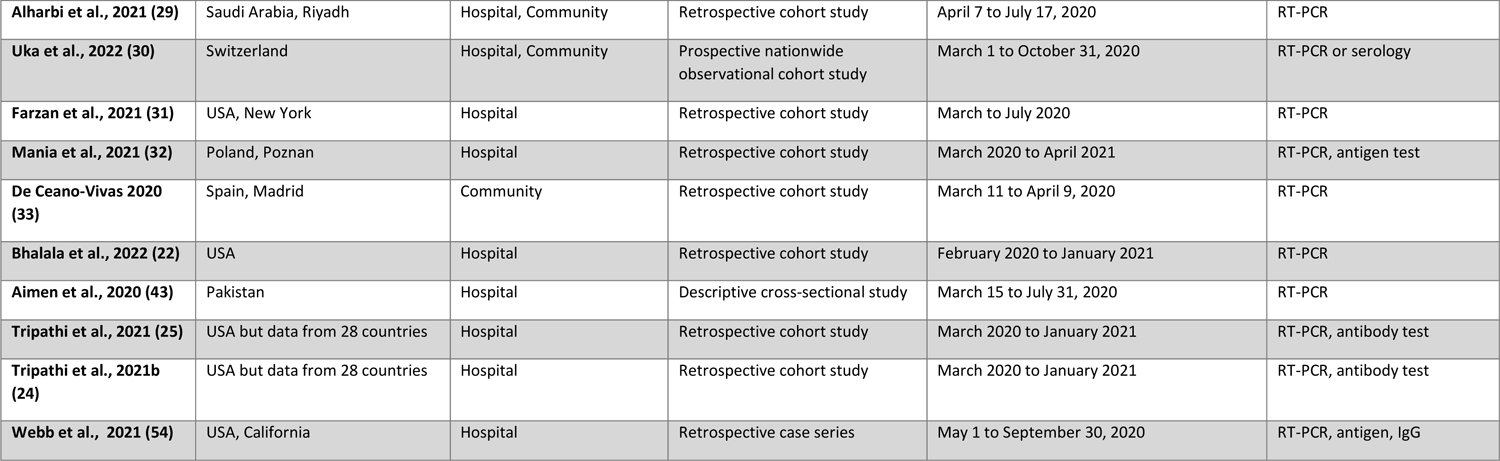
Methodological characteristics of included studies related to adverse outcomes of COVID-19 in children.

| FIRST AUTHOR,<br>YEAR OF PUBLICATION | COUNTRY, CITY | SETTING AT ENROLLMENT | STUDY DESIGN | RECRUITMENT DATES | COVID DIAGNOSIS |
| --- | --- | --- | --- | --- | --- |
| Shahid et al., 2021 (44) | Pakistan, Karachi | Hospital, community | Retrospective chart review | April to July 2020 | RT-PCR |
| Wang et al., 2020 (51) | China, Wuhan | Hospital | Retrospective case-control | January 25 to March 8, 2020 | RT-PCR |
| Drouin et al., 2021 (10) | Canada | Hospital | National prospective study | April 8 to December 31, 2020 | Microbiologically confirmed |
| Marques et al., 2021 (34) | Brazil, São Paulo | Hospital, community | Cross-sectional study | April 11, 2020 to April 22, 2021 | RT-PCR, serological tests |
| Diorio et al., 2020 (35) | US, Philadelphia | Hospital | Cross-sectional study | April 3 to May 15, 2020 | RT-PCR, serological tests |
| Snowden et al., 2021 (36) | US, Arkansas | Hospital, community | Cross-sectional study | February 1 to August 30, 2020 | RT-PCR |
| Martin et al., 2022 (11) | US | Hospital, community | Prospective cohort study | up to September 24, 2021 | PCR, antigen or antibody test |
| Moeller et al., 2020 (37) | Within and outside of Europe | Hospital, community | ERS Survey/ Cross-sectional study | April and May 2020 | Unknown |
| Antunez-Montes 2021 (12) | Mexico, Colombia, Peru, Costa Rica, Brazil | Hospital | Multicenter cohort study | July 1 to August 11, 2020 | RT-PCR |
| Okur et al., 2021 (13) | Turkey, Denizli | Hospital | Retrospective study | March 18, 2020, to January 18, 2021 | RT-PCR |
| Ouldali et al., 2021 (9) | France | Hospital, Community | National prospective cohort | February 15 to June 1, 2020 | RT-PCR, CT |
| Korkmaz et al., 2020 (14) | Turkey, Bursa | Hospital, Community | Retrospective observational cohort study | March 5 to May 5, 2020 | RT-PCR |
| Beken et al., 2021 (38) | Turkey, Istanbul | Community (ER) | Cross-sectional study | March 15 to May 31, 2020 | RT-PCR, CT |
| Nino et al., 2021 (39) | US, Washington | Community | Cross-sectional study | March to June 2020 | RT-PCR |
| Goktug et al., 2021 (40) | Turkey, Ankara | Hospital | Cross-sectional study | March to December 2020 | RT-PCR |
| Kainth et al., 2020 (15) | US, New York | Hospital | Single-centre retrospective cohort study | January 23 to April 23, 2020 | RT-PCR |
| Kapoor et al., 2021 (45) | India | Hospital | Case record review | March to December 2020 | RT-PCR |
| Antoon et al., 2021 (5) | US | Hospital | Multicenter retrospective cohort study | April to September 2020 | ICD-10 codes U.071 and U.072 |
| **Chopra et al., 2021 (41)** | India, New Delhi | Hospital | Cross-sectional analysis | March to November 2020 | RT-PCR |
| **Gotzinger et al., 2020 (16)** | 25 European countries | Hospital | Multicentre cohort study | April 1 to April 24, 2020 | RT-PCR |
| **Rao et al., 2021 (46)** | India, Mumbai | Hospital | Retrospective medical record review | March 19 to August 7, 2020 | RT-PCR |
| **Khan et al., 2021 (17)** | US, Saint Louis | Hospital | Retrospective cohort study | March 1, 2020, to February 28, 2021 | RT-PCR |
| **Graff et al., 2021 (18)** | US, Colorado | Hospital, Community | Retrospective cohort study | March to July 2020 | RT-PCR |
| **Kim et al., 2021 (19)** | US | Hospital, Community | Retrospective analysis | January 1 to July 24, 2020 | Any positive test result |
| **Howard et al., 2021 (20)** | US | Hospital, Community | Prospective cohort study | March 13 to September 28, 2020 | RT-PCR |
| **Verma et al., 2021 (21)** | US, New York | Hospital | Multicenter retrospective cohort study | March 1 to May 10, 2020 | RT-PCR |
| **Martinez-Garcia et al., 2021 (42)** | Mexico, Culiacán, Sinaloa | Hospital | Retrospective cross-sectional study | April 1 to June 30, 2020 | RT-PCR |
| **Metbulut et al., 2021 (52)** | Turkey, Ankara | Hospital, Community | Retrospective, case-control study | March 11 to November 10, 2020 | RT-PCR |
| **Gaietto et al., 2022 (53)** | US, Pittsburg | Community | Nested case-control stud | March 11 to December 21, 2020 | RT-PCR |
| **Grewal et al., 2021 (47)** | US, Michigan | Hospital | Retrospective chart review | March to August 2020 | RT-PCR, antibody test |
| **Chao et al., 2020 (48)** | US, Montefiore | Hospital | Retrospective chart review | March 15 to April 13, 2020 | RT-PCR |
| **Bhumbra et al., 2020 (23)** | US, Indianapolis | Hospital | Observational cohort study | up to May 4, 2020 | RT-PCR |
| **Funk et al., 2022 (26)** | Argentina, Australia, Canada, Costa Rica, Italy, New Zealand, Paraguay, Singapore, Spain, and the US | Community | Prospective cohort study | March 2020 to June 2021 | RT-PCR |
| **Oliveira et al., 2021 (49)** | Brazil | Hospital | Retrospective chart review | Feb 16, 2020 to January 9, 2020 | RT-PCR |
| **Bhavsar et al., 2021 (50)** | US, New Jersey | Hospital | Retrospective chart review | March 1 to May 31, 2020 | RT-PCR, serum IgG |
| **Fisler et al., 2020 (27)** | US, New York | Hospital | Retrospective cohort study | February 1 to April 24, 2020 | RT-PCR |
| **Alfraij et al., 2021 (28)** | Kuwait and Saudi Arabia | Hospital | Multicenter, retrospective, cohort study | March 1 to August 1, 2020 | RT-PCR |
| **Alharbi et al., 2021 (29)** | Saudi Arabia, Riyadh | Hospital, Community | Retrospective cohort study | April 7 to July 17, 2020 | RT-PCR |
| **Uka et al., 2022 (30)** | Switzerland | Hospital, Community | Prospective nationwide observational cohort study | March 1 to October 31, 2020 | RT-PCR or serology |
| **Farzan et al., 2021 (31)** | USA, New York | Hospital | Retrospective cohort study | March to July 2020 | RT-PCR |
| **Mania et al., 2021 (32)** | Poland, Poznan | Hospital | Retrospective cohort study | March 2020 to April 2021 | RT-PCR, antigen test |
| **De Ceano-Vivas 2020 (33)** | Spain, Madrid | Community | Retrospective cohort study | March 11 to April 9, 2020 | RT-PCR |
| **Bhalala et al., 2022 (22)** | USA | Hospital | Retrospective cohort study | February 2020 to January 2021 | RT-PCR |
| **Aimen et al., 2020 (43)** | Pakistan | Hospital | Descriptive cross-sectional study | March 15 to July 31, 2020 | RT-PCR |
| **Tripathi et al., 2021 (25)** | USA but data from 28 countries | Hospital | Retrospective cohort study | March 2020 to January 2021 | RT-PCR, antibody test |
| **Tripathi et al., 2021b (24)** | USA but data from 28 countries | Hospital | Retrospective cohort study | March 2020 to January 2021 | RT-PCR, antibody test |
| **Webb et al., 2021 (54)** | USA, California | Hospital | Retrospective case series | May 1 to September 30, 2020 | RT-PCR, antigen, IgG |

As per our inclusion criteria, the studies had no geographical limitations. Twenty studies were conducted in the United States (US) (5, 11, 15, 17–23, 27, 31, 35, 36, 39, 47, 48, 50, 53, 54), two in Brazil (34, 49), and one in Canada (10). In addition, five studies took place in Europe, of which one in France (9), one in Switzerland (30), one in Spain (33), one in Poland (32) and one across 25 European countries (16). There were also five studies from Turkey (13, 14, 38, 40, 52), three studies from India (41, 45, 46), two from Pakistan (43, 44), two from Saudi Arabia (28, 29) and one from China (51). Finally, six studies combined data from across countries (12, 24–26, 37, 42). The diagnosis of COVID-19 was primarily performed with a PCR test, except for three studies where the diagnosis was made with the ICD Classification (5), was microbiologically confirmed (10) or was unknown (37).

### Hospital admission among children recruited from the community setting

As noted in **Figure 2**, among children recruited from within the community setting (as a proxy of disease severity), neonates had higher odds of being hospitalised due to SARS-CoV-2 infection compared to older children (OR: 14.7; 95% CI: 6.8-31.8, I^2^=0%, high certainty). Similarly, infants under the age of 3 months (including neonates) were also more likely to be hospitalised (OR=7.9; 95% CI: 2.8-22.2, I^2^=0%, low certainty). Concerning BMI, underweight children (OR= 3.9, 95% CI: 2.1-7.1, I^2^=0%, moderate certainty) or severely obese (OR= 4.8, 95% CI: 1.9-12.1, I^2^=0%, low certainty) had higher odds for hospital admission. The presence of metabolic disorders also increased the odds of hospital admissions of SARS-CoV-2 infected children (OR= 29.6, 95% CI: 3.0-288.1, I^2^=0%, high certainty).

**Figure 2.**
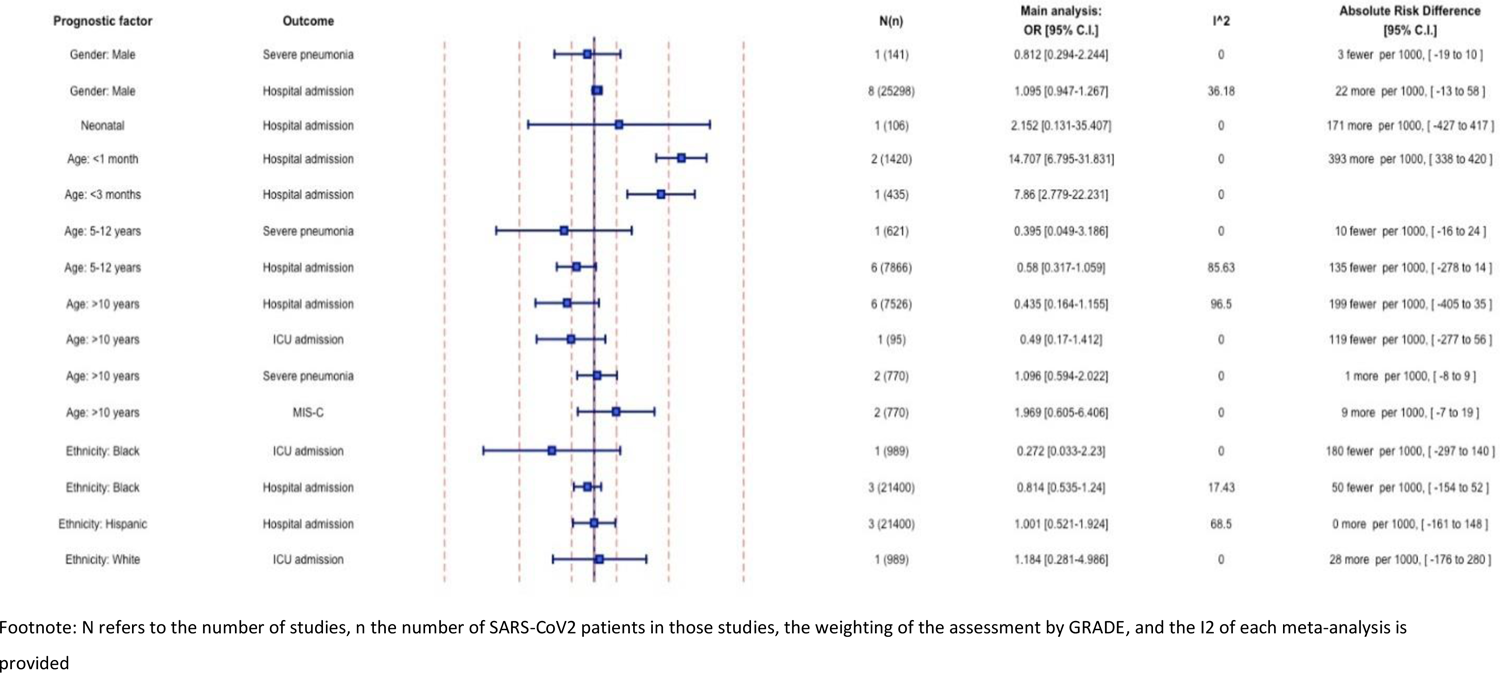
Demographic characteristics and COVID-19 severity outcomes among children recruited from the community setting.

Concerning comorbidities (**Figure 3**), having at least one underlying disease was found to increase the odds of hospitalisation by 3.2 times (95% CI: 2.3-4.6, I^2^=58.8%, low certainty). Our meta-analyses indicated that children with underlying cardiovascular diseases were approximately five times more likely to be hospitalised (OR = 5.0, 95% CI: 4.3-5.8, I^2^ = 0%, moderate certainty), followed by those with respiratory disorders (OR =3.5, 95% CI: 2.5-4.9, I^2^=20.4%, moderate certainty). Children with neuromuscular disorders were 3.2 times more likely to be hospitalised due to COVID-19, while the hospital admissions of those with neurologic conditions exceeded by 236 per 1,000 infections (95% CI: 78-336, I^2^=0%, moderate certainty) the respective admissions of COVID-19 paediatric patients without any neurologic condition (OR= 3.1, 95% CI: 1.4-6.9, I^2^ =0%, moderate certainty). Elevated odds were additionally calculated for hospital admission with regards to pre-existing malignancies (OR=14.8, 95% CI: 2.1-106.5, I^2^ = 62.5%, low certainty), kidney disease (OR=9.5, 95% CI: 1.0-88.3, I^2^=0%, very low certainty) and haemato- immunological diseases (OR=6.3, 95% CI: 2.2-18.1, I^2^=67.2%, low certainty).

**Figure 3.**
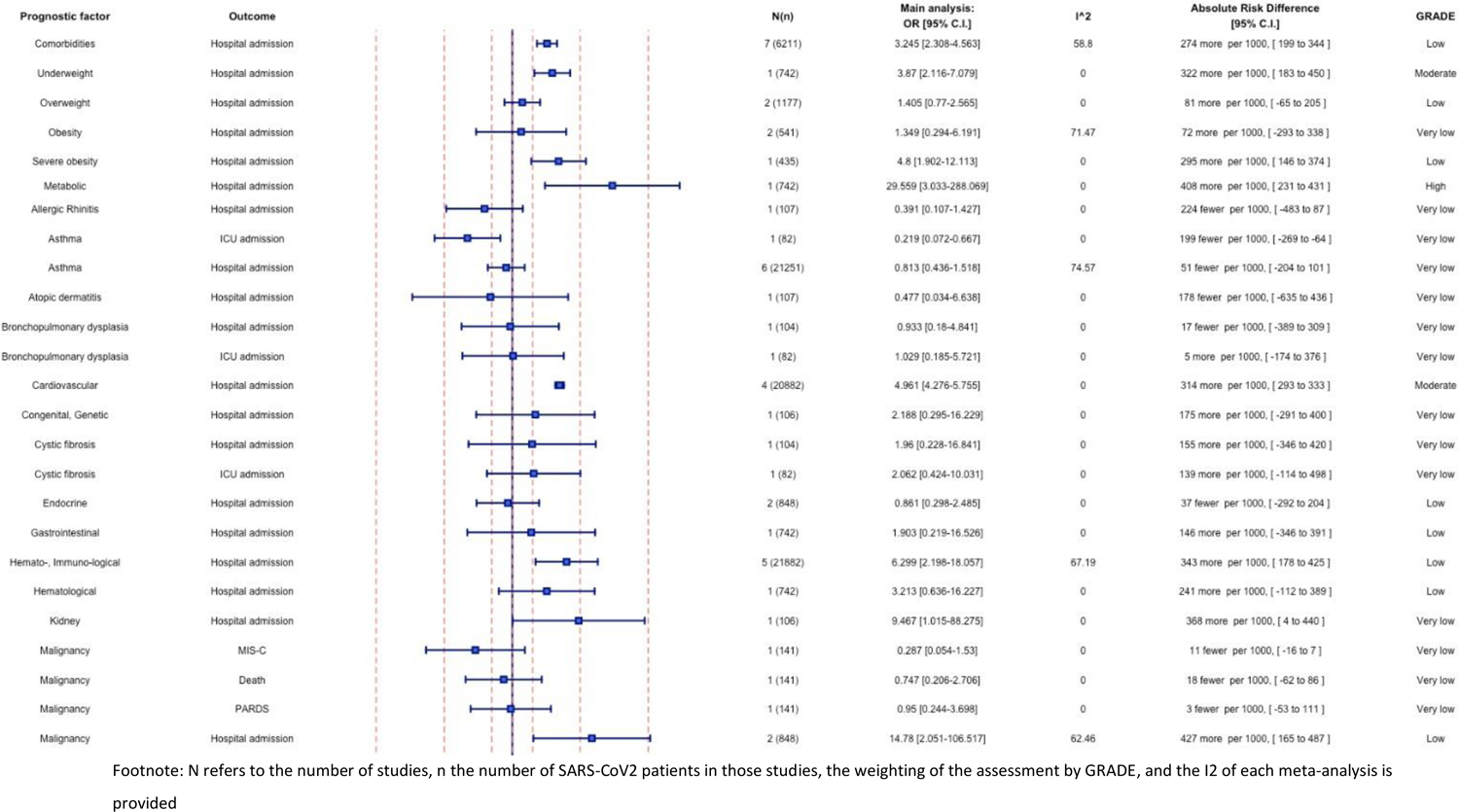
Underlying comorbidities and COVID-19 severity outcomes among children recruited from the community setting.

### Mechanical ventilation, multisystem inflammatory syndrome in children (MIS-C), ICU admission and death among hospitalised children

Among the demographic characteristics of hospitalised paediatric patients presented in **Figure 4**, male gender (OR=1.3, 95% CI: 1.9-1.5, I^2^=0%, moderate certainty) and black race (OR=1.3, 95% CI: 1.1-1.5, I^2^=0%, moderate certainty) were associated with increased odds for the need of mechanical ventilation. Also, ICU admission was 1.5 times higher in black children (95% CI: 1.1-1.9, I^2^=13%, moderate certainty). Regarding MIS-C development following SARS-CoV-2 infection, higher odds were estimated among those of black race (OR=1.3, 95% CI: 1.0-1.7, I^2^=13%, moderate certainty), and children aged between 5-12 years (OR=1.8, 95% CI: 1.0-3.3, I^2^=0%, low certainty) or over 10 years (OR=2.5; 95% CI: 1.5-4.14, I^2^=0%, low certainty). Neonates had lower odds of developing MIS-C (OR=0.3; 95% CI: 0.1-0.9, I^2^=0%, low certainty) compared to children of older ages.

**Figure 4.**
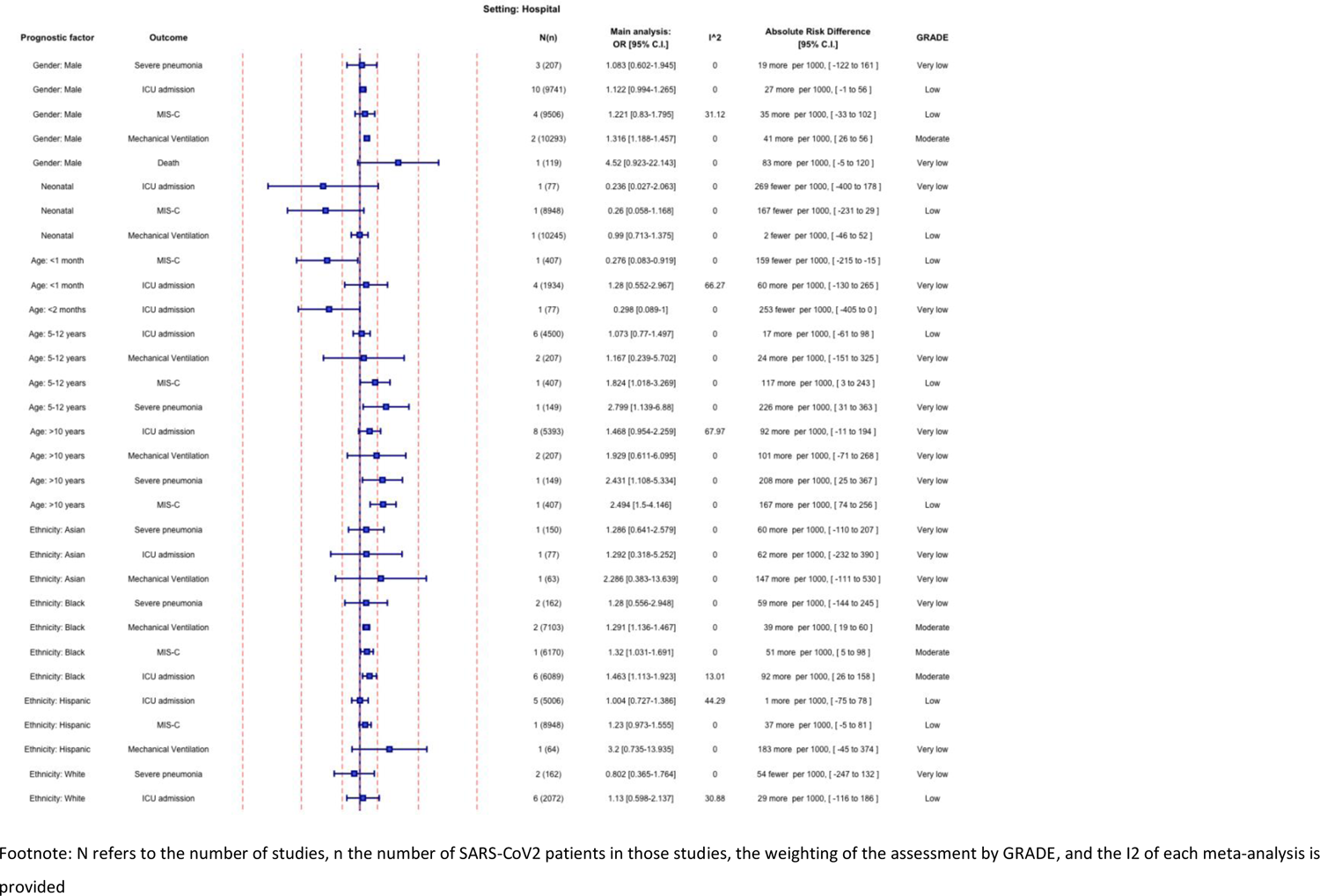
Demographic characteristics and COVID-19 severity outcomes among hospitalised paediatric patients.

Concerning underlying comorbidities (**Figure 5**), paediatric hospitalised patients with pre-existing conditions had higher odds (OR=2.5; 95% CI: 1.6-3.7, I^2^=67%, low certainty) of being admitted to ICU, while no statistically significant association was found for the need for mechanical ventilation and MIS-C development. The highest OR was estimated for children with congenital/genetic disease (OR=7.4, 95% CI: 2.4-22.7, I^2^=56.5%, low certainty), while obesity was associated with an elevated likelihood both for ICU admission (OR=2, 95% CI: 1.2-3.4, I^2^=59.1%, low certainty) and mechanical ventilation (OR=1.2, 95% CI: 1.0-1.4, I^2^=13%, moderate certainty) in hospitalised paediatric patients. Moreover, children with cancer were 60% (OR=1.6, 95% CI: 1.2-2.4, I^2^=0%, moderate certainty) and 150% (OR=2.5, 95% CI: 1.2-5.5, I^2^=0%, moderate certainty) more likely to be mechanically ventilated and admitted to ICU, respectively. Cardiovascular diseases were also identified as factors associated with both mechanical ventilation (OR=1.6, 95% CI: 1.3-2, I^2^=0%, moderate certainty) and ICU admission (OR=3.1, 95% CI: 2.5-3.7, I^2^=0%, moderate certainty). Finally, the pre-existence of any respiratory disease was associated with 2.7 higher odds of ICU admission (95% CI: 2-3.7, I^2^=0%, moderate certainty).

**Figure 5.**
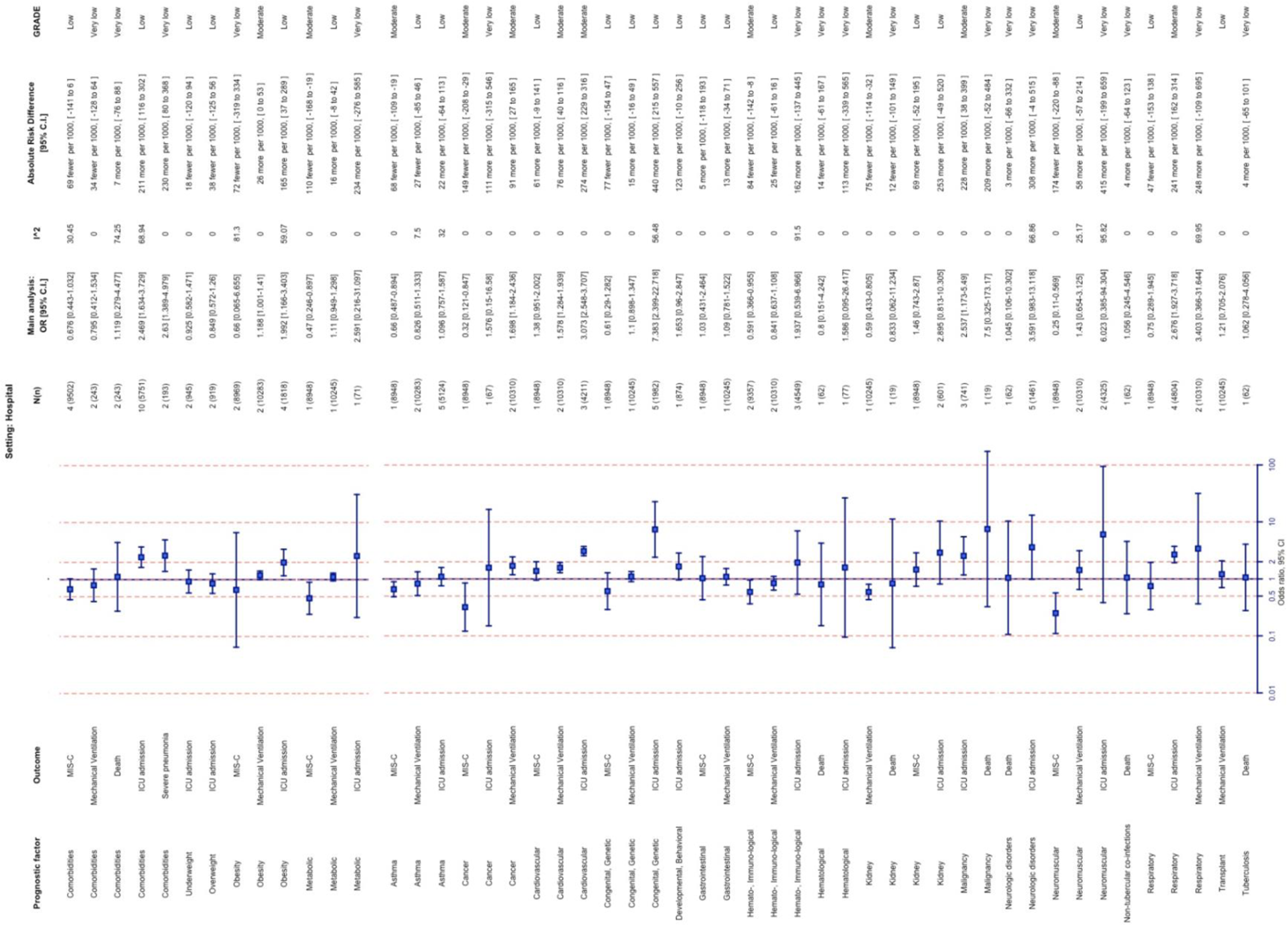
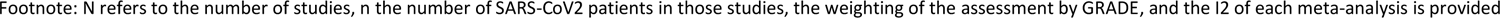
Underlying comorbidities and COVID-19 severity outcomes among hospitalised paediatric patients.

In contrast with the abovementioned findings, which show an increased risk for adverse COVID-19 outcomes, comorbidities seemed to be associated with lower odds for MIS-C development after SARS-CoV-2 infection, within the context of this review and for the timepoint of follow-up that was performed in the original studies. Children with a metabolic disorder were estimated to have approximately 50% smaller odds of developing MIS-C (OR=0.5, 95% CI: 0.2-0.9, I^2^=0%, moderate certainty). Likewise, neuromuscular disorders (OR=0.3, 95% CI: 0.1-0.6-1.4, I^2^=0%, moderate certainty), haemato-immunological disorders OR=0.6, 95% CI: 0.4-0.95, I^2^ =0%, moderate certainty), cancer (OR=0.3, 95% CI: 0.1-0.8, I^2^=0%, moderate certainty) and asthma (OR=0.7, 95% CI: 0.5-0.6, I^2^=0%, moderate certainty) were also related to a decreased likelihood of MIS-C occurrence after SARS-CoV-2 infection. Finally, paediatric patients with kidney diseases had lower odds of being mechanically ventilated (OR=0.6, 95% CI: 0.4-0.6, I^2^=0%, moderate certainty). We did not identify any demographic factors or comorbidities statistically significantly associated with death among hospitalised paediatric patients with COVID-19.

## DISCUSSION

Our work reflects an extensive body of research on the risk factors linked to COVID-19 progression in paediatric patients. It is noteworthy that, within our study, we were able to use recruiting settings as a proxy for disease severity to divide study populations. In addition, we examined the COVID-19 endpoints separately, including hospital admission, ICU admission, mechanical ventilation, MIS-C, and death, as opposed to prior studies that aggregated these outcomes under the heading of severe COVID-19 progression.

According to the results of this comprehensive analysis, children and adolescents had higher odds of hospitalisation and ICU admission if they had specific demographic characteristics and concomitant conditions. Corroborating studies also highlight that the odds of a severe COVID-19 outcome are substantially elevated for children with underlying risk factors compared with healthy children(55), underscoring that two or more comorbidities rather than single entities pose a risk for more severe courses of SARS-CoV-2 infection in children (55, 56).

### Hospitalisation, ICU admission and mechanical ventilation

Our meta-analysis focusing on global data indicated that among children recruited within the community setting, those younger than three months and neonates were more likely to be hospitalised, a finding which potentially could be interpreted as a clinical practice of caution for these early-aged patients, more than an index of disease severity. The existence of co-morbidities, such as metabolic disorders, as well as cardiovascular, haemato-immunological, kidney, respiratory, neurological, neuromuscular disorders and cancer were found to significantly affect the likelihood of hospital admission. This is in line with previous research both internationally (57), as well as from Europe (58), which has shown that the adjusted odds of hospitalisation were higher among paediatric cases with at least one comorbidity compared to healthy individuals. An additional finding of this study emphasised the significance of obesity in the hospitalisation of COVID-19 paediatric patients, which has been previously identified as a risk factor for severe COVID-19 in both adults and children (59, 60). Possible explanations for this relationship include a decline in lung function, modifications to the microbiota, an increase in pro-inflammatory chemicals, and adjustments to the immune system (61), or simply this is a result of the application of the cautionary principle for this paediatric population. Concerning demographic characteristics, previous research has estimated that the likelihood of hospitalisation is highest in the youngest age groups in European countries, subsequently reduces with age until nine years, and then rises with each passing year from 12 to 17 years (58). However, the heightened odds of hospitalisation seen for children younger than two months may reflect lower thresholds for admission of infants and neonates (62). Therefore, hospitalisation indexes may not be an appropriate proxy for illness severity in this age group, and hence additional research is necessary to clarify this finding. Finally, literature originating from the US suggests that male gender and black/non-white ethnicity may also be risk factors for hospitalisation in COVID-19 paediatric patients (57, 58). However, these factors did not have a statistically significant association with hospitalisation in our meta-analysis.

With reference to ICU admission which could be noted as a core outcome of COVID-19 severity, only black ethnicity was significantly linked to elevated odds for COVID-19 paediatric patients among the demographic factors in our analysis, a factor which may be a proxy for other factors associated with the likelihood of hospitalisation, such as earlier access to healthcare etc. Previous systematic reviews and meta-analyses have indicated the effect of age (<1 month) and the male gender as risk factors for admission to the ICU (63). Our study demonstrated that the presence of comorbid disorders such as obesity, cancer, congenital/genetic, haemato-immune, respiratory, and cardiovascular diseases significantly increased the likelihood that COVID-19 paediatric patients would be admitted to ICU, a finding however which also applies to other viral infections and not only to SARS-CoV-2. In alignment with our findings, the pooled estimates presented in the review of Shi et al. (2021) showed that congenital heart disease, chronic pulmonary disease and obesity increased the odds of admission to ICU (63). As for mechanical ventilation, similarly to ICU admission, the statistically significant risk factors that were detected in our meta-analysis included male gender and black ethnicity, as well as obesity, cancer, and cardiovascular diseases.

### MIS-C

In our meta-analysis, the odds for MIS-C development in COVID-19 paediatric patients were found to be increased in children older than 5 years old and of Black ethnicity. On the contrary, children younger than 1 month and those with metabolic, haemato-immunological, and neuromuscular disorders, along with asthma and cancer were estimated to have lower odds of developing MIS-C. Our findings are supported by other systematic reviews with previous cut-off dates. According to the literature that is currently accessible, children who develop MIS-C are frequently previously healthy and typically in their early to middle childhood years. In the review of Rafferty et al. (64) ages ranged from 7 months to 20 years, with median ages ranging from 7 to 10 years. Additionally, despite some hypotheses that being overweight may enhance the probability of developing MIS-C, most studies reported little to no association (65). In contrast to an injury brought on by an acute SARS-CoV-2 infection, the recently described paediatric MIS appears to be the outcome of a chronic immune response (66). Moreover, as there is a lag time between SARS-CoV-2 infection and the development of MIS-C, it is possible that the follow-up time within the included studies may have been insufficient to catch all cases of MIS-C, hence, more research is required to draw firm conclusions.

Numerous research studies have suggested that African American, African/Afro-Caribbean, and Hispanic children may be disproportionately impacted by MIS-C in terms of race and ethnicity – however, this disparity is likely a result of health disparities and access to healthcare or other societal factors. African/Afro-Caribbean children frequently made up the largest percentage of cases in European studies with race/ethnicity data, ranging from 38 percent to 62 percent of MIS-C patients (65). The overrepresentation of black and Hispanic racial/ethnic groups among MIS-C patients may partly be due to the greater SARS-CoV-2 infection rates among these groups within the earlier phases of the pandemic (68, 69). Still, others have postulated that genetic predisposition might play a role in susceptibility to MIS-C (70, 71), while racial and ethnic minority groups are also generally more likely to encounter hurdles to healthcare access, exacerbating the issue (72, 73).

### Death

Currently, given the small number of fatal cases in paediatric COVID-19 patients, the risk factors for mortality are scarce. In our study, no risk factors were significantly associated with increased odds of death. However, previous systematic reviews and meta-analyses demonstrated that underlying conditions are associated with increased odds of mortality, although estimated from very low-quality evidence. Additionally, age was appraised as a risk factor, with an increased likelihood among ages between four and ten years old (63). With regard to the role of comorbidities, in a recent review by Wijaya et al. (74), an increased risk for mortality was associated with malignancies, particularly haematological malignancies.

### Strengths and limitations

Only peer-reviewed studies are included in this systematic review and meta-analysis, and study populations were segmented using recruiting settings as a proxy for disease severity. In addition, unlike earlier studies that combined severity outcomes under the category of severe COVID-19 development, we looked at the COVID-19 endpoints separately, including hospital admission, ICU admission, mechanical ventilation, MIS-C, and death. However, the limitations of our study should also be acknowledged as our results may be affected by confounding as the original studies included in this meta-analysis did not control for confounding factors and presented unadjusted odds. Furthermore, the included studies reflect mainly the pre-vaccination era, while only a few included data up to mid-2021. Furthermore, the research identified within this review does not provide information about different SARS-CoV-2 variants. Additionally, we included studies with no geographical limitation, which made it difficult to compare the results. However, since the results have similarities with European data (58), European physicians and policymakers may also benefit from them. Furthermore, there was evidence of heterogeneity as the definition of specific prognostic factors, such as congenital disorders, obesity, and respiratory disorders, were ambiguous in some of the included studies and variability in the definitions may have also contributed to the observed variability. Given that a direct comparison among different risk factors was not performed and similar effect sizes were found, no conclusions can be drawn on the comparative ranking or cumulative/synergistic effect of specific prognostic factors leading to COVID-related adverse outcomes. Nevertheless, conclusions on the individual impact of specific comorbidities can be drawn.

## CONCLUSION

The findings of this systematic literature review and meta-analysis suggest that there were a number of factors associated with an increased likelihood of hospital, ICU admission, mechanical ventilation and MISC-2 in paediatric populations. It is unclear to which extent the increased hospital admission noted for younger children and for children with specific comorbidities (obesity, cancer, respiratory and cardiovascular disorders) were associated with disease severity or the application of precautionary clinical care for these higher-risk groups. It is notable however that these factors were also associated with ICU admission while certain comorbidities (obesity, cancer, and cardiovascular disease) were also associated with mechanical ventilation, an invasive procedure not easily classifiable as a preventative care measure, indicating that these factors also play a role in disease severity as similarly noted among adults (75). This evidence may assist policymakers and public health authorities in emergency preparedness planning in the post-pandemic COVID-19 era.

## Supporting information

search strategy

## Funding

European Centre for Disease Prevention and Control (ECDC) under specific contract No. 19 ECD.12254 within Framework contract ECDC/2019/001 Lot 1B.

## Declaration of interests

We declare no competing interests.

## Data sharing statement

Data sharing is not applicable to this article as no new data were created or analysed in this study.

## Data Availability

All data produced in the present work are contained in the manuscript

## Notes

### Competing Interest Statement

The authors have declared no competing interest.

