## Supplementary material for "Prognostic factors for mortality, ICU, MIS-C and hospital admission due to SARS-CoV-2 in paediatric patients: A systematic review and meta-analysis": search strategy

### APPENDIX 1 – Search Strategy

**Ovid MEDLINE(R) ALL <1946 to February 14, 2022>**

1 exp coronavirus/ or exp Coronavirus disease 2019/ or exp coronavirus infections/ 157801
2 Coronaviridae Infections/ or Coronaviridae/ or SARS-CoV-2/ or COVID-19/ 143620
3 (("2019" adj (novel or new) adj corona*) or ("2019" adj (CoV or nCoV)) or (coronavirus adj (disease adj "2019")) or COVID19 or COVID-19 or ((Novel or New) adj Corona*) or SARS2 or SARS-CoV-2 or (SARS adj2 (coronaviridae or coronavirus)) or ((sars or Coronavirus) adj "2") or nCov or 2019ncov or (severe adj acute adj respiratory adj syndrome) or SARs or Sars-cov or ((sars-associated or sars-related) adj (cov or coronavirus))).mp. 238966
4 Betacoronavirus 1/ or Betacoronavirus/ 33290
5 or/1-4 249740
6 exp cohort analysis/ 2296421
7 exp longitudinal study/ 155295
8 exp prospective study/ 615095
9 cohort*.tw. 729143
10 "controlled clinical trial".pt. 94698
11 or/6-10 2690715
12 exp mortality/ 414881
13 exp hospital mortality/ 47011
14 Death/ or death*.mp. 1021755
15 all-cause death.mp. 6648
16 all-cause mortality.mp. 43077
17 exp hospitalization/ 273986
18 exp patient admission/ 25933
19 exp patient readmission/ 21035
20 mortality.mp. 1292756
21 (died or fatal or decrease* or non-survivor or non-survival or survival).mp. 4103283
22 (recovered or discharge* or alive or non-fatal or nonfatal or survivor).mp. 576672
23 ("hospital stay*" or "patient readmission" or "patient admission" or utilisation or utilization or morbidity or "hospital care" or "hospital treatment").mp. 816108
24 exp tidal volume/ or LPVS.mp. or "pressure* adj3 limited*".mp. or "protective adj3 ventilliation".mp. or exp respiration, artificial/ or ALI.mp. or ARDS.mp. 110886
25 respiratory disease syndrome/ or adult respiratory distress syndrome/ or acute lung injury/ or acute respiratory distress syndrome/ or "tidal volume".mp. 46023
26 ((intensive or critical) adj1 care).mp. 247626
27 exp intensive care units/ or exp intensive care/ or ICU.mp. or exp critical care/ 181298
28 critical illness.mp. or exp critical illness/ 40709
29 ("mechanical ventillation" or "mechanical ventilation").mp. 51160
30 ITU.mp. 1029
31 intensive therapy unit.mp. 548
32 Systemic Inflammatory Response Syndrome/ 6671
33 ((multisystem* or multi-system*) and inflammat* and syndrome*).mp. 3083
34 or/12-33 6346617
35 (young* or infant* or baby or babies or pediatri* or paediatr* or newborn* or neonat*).mp. 3302667
36 Infant, Newborn/ or Infant/ 1204221
37 (toddler* or preschool* or child* or pediat* or paediat* or kid or kids or prepubescen* or prepuberty* or puberty or pubescen* or teen* or young* or youth* or minors* or under ag* or underag* or juvenile* or girl* or boy* or preadolesc* or adolesc* or nursery or prekindergarten or kindergarten* or early childhood education or preschool* or elementary education or elementary school* or primary education or primary school* or K-12* or K12 or 1st-grade* or first-grade* or grade 1 or grade one or 2nd-grade* or second-grade* or grade 2 or grade two or

3rd-grade* or third-grade* or grade 3 or grade three or 4th-grade* or fourth-grade* or grade 4 or grade four or 5th-grade* or fifth-grade* or grade 5 or grade five or 6th-grade* or sixth-grade* or grade 6 or grade six or intermediate general or middle school* or secondary education or secondary school*OR 7th-grade* or seventh-grade* or grade 7 or grade seven or 8th-grade* or eight-grade* or grade 8 or grade eight or 9th-grade* or ninth-grade* or grade 9 or grade nine or 10th-grade* or tenth-grade* or grade 10 or grade ten or 11th-grade* or eleventh-grade* or grade 11 or grade eleven or 12th-grade* or twelfth-grade* or grade 12 or grade twelve or junior high* or highschool* or high school* or preuniversity or pre-university or college* or tertiary education or tertiary school*OR postsecondary education or postsecondary school* or prevocational or vocational or classroom* or curricul* or education* or learner* or lesson* or pupil* or school* or student*).ti,ab,kf. 3682573
38 toddlers/ or preschool children/ or young children/ or children/ or pediatrics/ or preadolescents/ or youth/ or adolescents/ or early adolescents/ or late adolescents/ or nursery schools/ or kindergarten/ or early childhood education/ or preschool education/ or preschool teachers/ or elementary secondary education/ or grade 1/ or grade 2/ or grade 3/ or grade 4/ or grade 5/ or grade 6/ or grade 7/ or grade 8/ or grade 9/ or grade 10/ or grade 11/ or grade 12/ or elementary education/ or elementary schools/ or elementary school students/ or elementary school teachers/ or primary education/ or public schools/ or public school teachers/ or middle schools/ or middle school students/ or junior high schools/ or junior high school students/ or secondary education/ or secondary schools/ or secondary school students/ or secondary school teachers/ or high schools/ or high school students/ or college students/ or colleges/ or two year college students/ or two year college students/ or vocational education/ or vocational schools/ or students/ 3306537
39 or/35-375266221
40 5 and 34 and 39 17177
41 limit 40 to yr="2020 -Current" 16221
42 limit 40 to yr="2021 -Current" 11389
43 41 and 11 4073
44 42 and 11 2923

**Embase <1974 to 2022 February 11>**

1 exp coronavirus/ or exp Coronavirus disease 2019/ or exp coronavirus infections/ 228287
2 Coronaviridae Infections/ or Coronaviridae/ or SARS-CoV-2/ or COVID-19/ 65502
3 (("2019" adj (novel or new) adj corona*) or ("2019" adj (CoV or nCoV)) or (coronavirus adj (disease adj "2019")) or COVID19 or COVID-19 or ((Novel or New) adj Corona*) or SARS2 or SARS-CoV-2 or (SARS adj2 (coronaviridae or coronavirus)) or ((sars or Coronavirus) adj "2") or nCov or 2019ncov or (severe adj acute adj respiratory adj syndrome) or SARs or Sars-cov or ((sars-associated or sars-related) adj (cov or coronavirus))).mp. 260847
4 Betacoronavirus/ or Severe acute respiratory syndrome coronavirus 2/62058
5 or/1-4 271260
6 exp cohort analysis/ 805632
7 exp longitudinal study/ 167923
8 exp prospective study/ 744821
9 exp follow up/ 1797734
10 cohort*.tw. 1227355
11 or/6-10 3391287
12 exp mortality/ 1221437
13 exp hospital mortality/ 48075
14 death*.mp. 1547529
15 all-cause mortality.mp. 90249
16 all-cause death*.mp. 13053
17 exp hospitalization/ 443756
18 exp patient admission/ 233236
19 exp patient readmission/ 80523
20 exp death/ 765079

21 (mortality or died or fatal or decease* or non-survivor or "non survivor").mp. [mp=title, abstract, heading word, drug trade name, original title, device manufacturer, drug manufacturer, device trade name, keyword heading word, floating subheading word, candidate term word] 2107598
22 (recovered or discharge* or alive or non-fatal or nonfatal or survivor).mp. 968702
23 respiratory distress syndrome, adult/ 26483
24 ((acute adj lung adj injury) or (shock adj lung) or ARDS).mp. [mp=title, abstract, heading word, drug trade name, original title, device manufacturer, drug manufacturer, device trade name, keyword heading word, floating subheading word, candidate term word] 51099
25 ((acute or adult) and (respiratory adj distress)).mp. 81963
26 ((intensive or critical) adj1 care).mp. 431955
27 exp intensive care units/ 239264
28 exp intensive care/ 755681
29 exp critical care/ 755681
30 ("critical illness" or ICU or ITU or "intensive therapy unit").mp.176416
31 exp critical illness/ 32903
32 ("mechanical ventillation" or "mechanical ventilation").mp. 84228
33 systemic inflammatory response syndrome/ 13839
34 ((multisystem* or multi-system*) and inflammat* and syndrome*).mp. 4270
35 or/12-345003787
36 (young* or infant* or baby or babies or pediatri* or paediatr* or newborn* or neonat*).mp. 3199626
37 infant/ 643730
38 newborn/ 563566
39 (toddler* or preschool* or child* or pediat* or paediat* or kid or kids or prepubescen* or prepuberty* or puberty or pubescen* or teen* or young* or youth* or minors* or under ag* or underag* or juvenile* or girl* or boy* or preadolesc* or adolesc* or nursery or prekindergarten or kindergarten* or early childhood education or preschool* or elementary education or elementary school* or primary education or primary school* or K-12* or K12 or 1st-grade* or first-grade* or grade 1 or grade one or 2nd-grade* or second-grade* or grade 2 or grade two or 3rd-grade* or third-grade* or grade 3 or grade three or 4th-grade* or fourth-grade* or grade 4 or grade four or 5th-grade* or fifth-grade* or grade 5 or grade five or 6th-grade* or sixth-grade* or grade 6 or grade six or intermediate general or middle school* or secondary education or secondary school* or 7th-grade* or seventh-grade* or grade 7 or grade seven or 8th-grade* or eight-grade* or grade 8 or grade eight or 9th-grade* or ninth-grade* or grade 9 or grade nine or 10th-grade* or tenth-grade* or grade 10 or grade ten or 11th-grade* or eleventh-grade* or grade 11 or grade eleven or 12th-grade* or twelfth-grade* or grade 12 or grade twelve or junior high* or highschool* or high school* or preuniversity or pre-university or college* or tertiary education or tertiary school* or postsecondary education or postsecondary school* or prevocational or vocational or classroom* or curricul* or education* or learner* or lesson* or pupil* or school* or student*).ti,ab,kf. 4754660
40 toddler/ or preschool child/ or child/ or pediatrics/ or juvenile/ or adolescent/ or nursery school/ or kindergarten/ or preschool child/ or primary education/ or secondary education/ or tertiary education/ or school child/ or middle school student/ or high school student/ or primary school/ or high school/ or high school graduate/ or school/ or college/ or college student/ 3214676
41 or/36-406804496
42 5 and 35 and 41 21530
43 limit 42 to yr="2020 -Current" 20211
44 limit 42 to yr="2021 -Current" 13762
45 43 and 11 5247
46 44 and 11 3885
